# Parent-of-Origin-Aware Genomic Analysis to Enhance Cascade Testing in Pancreatic Cancer: A Population-Based Study

**DOI:** 10.64898/2026.10.30.26366356

**Authors:** Phoebe T.M. Cheng, Carol Cremin, Lilian Cordova, Vahid Akbari, Tiffany Y. Leung, Eugene Cheung, Angela Bedard, Quan Hong, Ana Fisic, Jennifer Nuk, Mary-Jill Asrat, Angela Inglis, Allison Mindlin, Melanie O’Loughlin, Katherine Dixon, Kieran O’Neill, Alexandra T. Roston, Chuyi Zheng, Alshanee Sharma, Yaoqing Shen, Janine Senz, Yanni Wang, Daniel D. Chan, Alexandra Fok, Adam Kahnamelli, Steve Bilobram, Simon Chan, Robin Coope, Hyun-Wu Lee, Yongjun Zhao, Miruna Bala, Karen Mungall, Andy Mungall, Richard Moore, Dean A. Regier, Alice Virani, Fabio Feldman, Janessa Laskin, Marco Marra, Sophie Sun, Stephen Yip, Peter M. Lansdorp, Steven Jones, David F. Schaeffer, Daniel J Renouf, Kasmintan A. Schrader

**Affiliations:** Department of Medical Oncology, BC Cancer, Vancouver, Canada; Department of Medical Genetics, University of British Columbia, Vancouver, Canada; Canada’s Michael Smith Genome Sciences Centre, BC Cancer, Vancouver, Canada; Hereditary Cancer Program, BC Cancer, Vancouver, Canada; Terry Fox Laboratory, BC Cancer, Vancouver, Canada; Division of Genome Diagnostics, BC Children’s and Women’s Hospital, Vancouver, Canada; Department of Basic and Translational Research, BC Cancer Research Institute, Vancouver, Canada; School of Population and Public Health, University of British Columbia, Vancouver, Canada; Population Health Sciences, BC Cancer, Vancouver, Canada; Ethics and Spiritual Care, Provincial Health Services Authority, Vancouver, Canada; Pathology and Laboratory Medicine, University of British Columbia, Vancouver, Canada; Pancreas Centre BC, Vancouver, Canada; Cancer Genetics and Genomics Lab, BC Cancer, Vancouver, Canada

**Keywords:** pancreatic cancer, germline genetic testing, cascade testing, cancer prevention

## Abstract

Identifying germline pathogenic variants has therapeutic value for pancreatic ductal adenocarcinoma (PDAC) patients and critical implications for cascade genetic testing in families, the latter being an increasing focus for public health genomic initiatives. We measured cascade testing rates for one hundred consecutive hereditary PDAC probands (March 2014-December 2023) and assessed the potential impact of using a novel technology, Parent-of-Origin-Aware genomic analysis (POAga) to enhance cascade testing. POAga requires a new blood sample and enables variant segregation to the maternal or paternal lineage. Thirteen probands were successfully contacted about POAga and nine provided a research blood sample. Parental origin was determined for all variants, and if applied at the time of diagnosis, would have immediately refined risk estimates for at least 188 relatives. POAga may overcome a major barrier to cascade testing in PDAC by clarifying which side of the family is at risk, improving cascade testing effectiveness and familial cancer prevention strategies.

**What is known about the topic:** Despite significant expansion in universal genetic testing criteria, cascade testing rates in relatives remain sub-optimal and require novel strategies in order to realize the full preventive value of hereditary cancer identification.

**What this paper adds:** This study describes the real-world impact of a new technological advance, Parent-of-Origin-Aware genomic analysis, that fundamentally changes genetic risk assessment by identifying which parental branch is most likely to harbour risk. In doing so, it immediately focuses genetic counselling efforts on one side of the family, with important implications for patients, families, and health-system implementation.

## Introduction

Realization of the broad benefits of universal germline testing in newly diagnosed cancer patients relies on maximizing uptake of cascade genetic testing and access to effective cancer prevention and early detection in relatives once a hereditary cancer susceptibility has been identified. A key focus of the genetic counselling profession is to lead and support efforts to improve suboptimal cascade testing rates that hover around 30% in most studies, including clinic-mediated direct contact of relatives. (1-6)

In pancreatic ductal adenocarcinoma (PDAC), universal germline testing is recommended because germline pathogenic variants (PV) are identified in approximately 10-15% of patients and have important implications for therapy and familial cancer risk stratification through cascade testing.(7) Genetic counselling and cascade testing in these families is further complicated by the limited life expectancy of affected individuals, the short timeframe for sharing results with relatives, and the frequent inability to determine the parental origin of a variant, since most individuals with pancreatic cancer are diagnosed later in life and many of their parents are deceased.

Parent-of-Origin-Aware genomic analysis (POAga) is a novel long-read and single-cell template strand sequencing-based method that infers the parental origin of any variant without requiring parental samples. This technology has the potential to improve cascade testing and risk communication by enabling more precise estimates of familial cancer risk.

This study describes germline findings and cascade testing patterns in a population-based PDAC cohort in British Columbia (BC), and explores the pilot application of POAga to refine familial risk assessment and risk communication.

## Methods

### Cohort Identification

All individuals in BC diagnosed with PDAC between March 2014 and December 2023 who were consecutively identified to harbour PV/LPV in established pancreatic cancer susceptibility genes were identified through the Hereditary Cancer Program (HCP). Currently established pancreatic cancer susceptibility genes included: *ATM, BRCA1, BRCA2, CDKN2A, MLH1, MSH2, MSH6, PMS2, EPCAM, PALB2, STK11, TP53* as well as several genes associated with hereditary pancreatitis (5).

### Data Collection

Demographics, family histories, and germline results were extracted from HCP records. Clinical outcomes were assessed as well as cascade testing rates and impact of POAga on a subset of patients. Follow up data was collected up to May 2024. All diagnoses were confirmed by pathology review.

### Statistical analysis

Overall survival (OS) was calculated from the dates of diagnosis to last follow-up or death from any cause. Survival endpoints were estimated using the Kaplan-Meier method. Comparisons were made using the χ2 test.

### Cascade Testing Assessment

Cascade testing rates among first- and second-degree relatives were obtained through family history review using the provincial Progeny database. Uptake was cross-verified by chart review and patient reports.

### Parent-of-Origin–Aware Genomic Analysis

POAga combines two main technologies and is described in detail by Akbari *et al*. (9) Herein, Oxford Nanopore technology long-read sequencing identified variants and allele- specific methylation signatures at imprinted differentially methylated regions (iDMRs). In parallel, single-cell Strand-seq, was used to generate chromosome-length haplotypes. Subsequent parent-of-origin assignment was achieved by phasing long reads to the Strand-seq scaffold to infer parent-of-origin of each haplotype, and every variant along it, based on known maternal and paternal iDMRs. Combined, these resolved maternal vs. paternal origin of the previously known PV without the need for parental blood samples.

All patients provided informed written consent for genetic testing and had their clinical result and POAga research result disclosed to them by a certified genetic counsellor. Ethics approval for this study was obtained from the BC Cancer Research Ethics Board (UBC BC Cancer REB), Reference Numbers: H18-03697 and H23-00699.

## Results

### Cohort Characteristics and Oncologic Outcomes

One hundred probands with PV/LPV were identified (Table 1). Variants occurred most frequently in homologous recombination repair (HRR) genes (n=47; 9 *BRCA1*, 35 *BRCA2*, 3 *PALB2*), followed by *ATM* (n=28), mismatch repair (MMR) genes (n=13), *CDKN2A* (n=11), and *TP53* (n=1). Notably, 47% of probands passed away within one year of receiving their genetic test result.

**Table 1.** Patient and disease characteristics of PDAC patients with PGVs (n=100). HRR= homologous recombination repair, MMR= mismatch repair, FDR= first-degree relative, FPC= familial pancreatic cancer, *Includes three double heterozygotes in *BRCA2/ATM* (n=2), and *BRCA2/MUTYH* (n=1), **Includes one double heterozygote with both an MMR and targetable HRR gene (PALB2/MSH6), ***Includes one double heterozygote in *PMS2/MITF*, ****Includes two double heterozygotes each in *ATM/APC* and *ATM/MUTYH*

| Characteristic | All Patients, N (%) |
|---|---|
| **Age at PDAC diagnosis, years (Median, range)** | 66 (36-89) |
| **Sex assigned at birth** |  |
| Female | 48 (48) |
| Male | 52 (52) |
| **Targetable HRR genes** | 47 (47) |
| *BRCA1* | 9 (9) |
| *BRCA2\** | 35 (35) |
| *PALB2\*\** | 3 (3) |
| **MMR genes** | 13 (13) |
| *MSH6* | 5 (5) |
| *MSH2* | 4 (4) |
| *PMS2\*\*\** | 4 (4) |
| **Other genes** |  |
| *ATM\*\*\*\** | 28 (28) |
| *CDKN2A* | 11 (11) |
| *TP53* | 1 (1) |
| **Mainstream Testing initiated by non-genetics provider** | 17 (17) |
| **Ethnicity** |  |
| European | 59 (59) |
| East Asian | 14 (14) |
| South/Central Asian | 4 (4) |
| Ashkenazi Jewish | 2 (2) |
| African | 1 (1) |
| Middle Eastern | 1 (1) |
| Mixed - European/Indigenous | 2 (2) |
| Mixed - European/Ashkenazi Jewish | 1 (1) |
| Ethnicity not reported | 16 (16) |
| **Risk factors** |  |
| Smoking history | 51 (51) |
| Pre-existing diabetes | 14 (14) |
| Family history of pancreatic cancer | 26 (26) |
| Chronic pancreatitis (pathology) | 8 (8) |
| **Personal history of another cancer** | 39 (39) |
| **Affected FDR (FPC, Reported)** | 18 (18) |
| **Performance status** |  |
| ECOG 0 or 1 | 69 (69) |
| ECOG > 1 | 23 (23) |
| ECOG not documented | 8 (8) |
| **Stage at diagnosis** |  |
| Upfront resectable | 27 (27) |
| Locally advanced/borderline resectable | 27 (27) |
| Metastatic | 46 (46) |
| Treatment outcomes (unresectable/metastatic) | Median OS (months) |
| All (n=62) | 17.6 |
| Targetable HRR genes (n=27) | 17.9 |
| *ATM (n=19)* | 17.9 |
| MMR genes (n=10) | 17.6 |
| *CDKN2A/TP53* (n=6) | 13.8 |

Most patients (73%) presented with unresectable disease. With median follow up of 24.9 months in living patients, median OS was 21.4 months in all 89 treated patients and 17.6 months in 62 upfront unresectable (metastatic or locally advanced) cases (Table 1). There was no difference between gene subgroups in stage distribution, or in the proportion of resectable versus upfront unresectable disease (p=0.39) (Table 1).

### Cascade Testing Uptake

During the study period, communication of cascade testing eligibility was via patient- mediated contact with their at-risk relatives. Among 641 first-degree relatives, 149 (23.2%) underwent testing; this rate was higher if only siblings and children were included at 30.8% (n=441). Among 1434 second-degree relatives, only 60 (4.2%) were tested. Among grandchildren, nieces and nephews, 8.76% (37/422) were tested, yielding a 43.2% mutation detection rate and 3.8% (16/422) overall hereditary cancer identification rate. If limited to parental extended SDRs, 2.27% (23/1012) were tested, yielding an 82.6% (19/23) mutation detection rate and 1.9% (19/1012) overall hereditary cancer identification rate.

Although there was sharing of information in 70% of families with at least one relative accessing cascade testing, the majority of at-risk relatives did not have testing. Across the whole cohort, an average 1.49 first-degree relatives were tested per index patient. Among the 70% of families where cascade testing occurred, an average of 2.12 first-degree relatives were tested per family.

Despite the 10-year study period, segregation of the PV was unknown in the majority of families (79%) and 75% of probands had no living parent, precluding parental cascade testing to clarify the side of the family at risk.

### Impact of POAga on Familial Risk Assessment

In February 2024, the full cohort was reviewed for possible POAga enrollment and a research blood draw. Medical records indicated that at least 70% of patients had passed away. Of the 13 patients who were successfully contacted, nine consented and provided a new blood sample (3 *ATM*, 2 *BRCA2*, 2 *MSH6*, 1 *BRCA1*, 1 *CDKN2A*) and four declined (2 *ATM*, 1 *ATM/MUTYH*, 1 *PALB2*), conferring a 69% POAga uptake rate. Table 2 shows POAga assigned parent-of-origin in all nine cases, allowing for a refined estimation of risks for extended relatives.

**Table 2.** Parent-of-Origin-Aware Genomic Analysis Results. SAAB= sex assigned at birth, PDAC= pancreatic ductal adenocarcinoma, Dx= diagnosis, FT=fallopian tube, CRC=colorectal cancer, PofO= Parent of Origin, POAga= Parent-of-Origin Aware Genomic Analysis

| POAga ID | SAAB | Age Group | Ancestry | Age at PDAC Dx | Other Cancer Site (age) | Type of Testing | Gene | Chr | Variant | Transcript | Mutation Type | Segregation | PofO Assigned | POAga Prediction | Parental Vital Status | Prediction confirmed by clinical cascade testing in relatives |
|---|---|---|---|---|---|---|---|---|---|---|---|---|---|---|---|---|
| POA-1 | M | 46-50 | European | 46-50 |  | Panel | *BRCA2* | 13 | c.5350_5351delAA (p.Asn1784Hisfs*2) | NM_000059 | Indel | Unknown | Yes | Paternal | Deceased | *Pending* |
| POA-2 | F | 56-60 | European | 51-55 | FT (56-60) | Panel | *BRCA2* | 13 | c.6070C>T (p.Gln2024*) | NM_000059 | SNV | Known | Yes | Paternal | Living (Mat)<sup>b</sup> | Yes |
| POA-3 | M | 76-80 | European | 71-75 |  | Panel | *BRCA1* | 17 | c.5117G>A (p.Gly1706Glu)<sup>a</sup> | NM_007294.3 | SNV | Unknown | Yes | Paternal | Deceased | *Pending* |
| POA-4 | F | 56-60 | Asian | 51-55 | Breast (31-35) | Panel | *ATM* | 11 | c.5679_5691del13 (p.E1894Dfs*19) | NM_000051.3 | Indel | Unknown | Yes | Maternal | Living | *Pending* |
| POA-5 | M | 71-75 | European | 66-70 |  | Panel | *CDKN2A* | 9 | c.202G>C (p.A68P) | NM_000077.4 | SNV | Unknown | Yes | Maternal | Deceased | *Pending* |
| POA-6 | F | 76-80 | European | 71-75 | CRC (71-75) | Panel | *ATM* | 11 | c.5496+645T>G | NM_000051.3 | SNV | Unknown | Yes | Maternal | Deceased | *Pending* |
| POA-7 | M | 81-85 | European | 76-80 | Skin (46-50) | Panel | *MSH6* | 2 | c.3013C>T (p.Arg1005*) | NM_000179.2 | SNV | Known | Yes | Maternal | Deceased | Yes |
| POA-8 | F | 76-80 | European | 71-75 | Uterine (61-65) | Panel | *MSH6* | 2 | Del exon 5-10 | NM_000179.2 | Structural | Unknown | *Yes* | Paternal | Deceased | *Pending* |
| POA-9 | F | 76-80 | Unknown | 66-70 |  | Panel | *ATM* | 11 | c.4098_4099delTG (p.Cys1366*) | NM_000051 | Indel | Unknown | *Yes* | Maternal | Deceased | *Pending* |

<sup>a</sup>Founder mutation population = Spain
<sup>b</sup>Only maternal parent is still living

Figure 1 shows the numbers of at-risk maternal and paternal relatives (n=1192) for each of the 100 probands, illustrating the impact of POAga on genetic risk assessment if routinely performed for all probands. Among the 9 families where samples were available for POAga, at least 188 maternal and paternal relatives were initially considered at risk of carrying familial variants. As a result, if POAga had been immediately applied, genetic risks were doubled for 110 relatives based on the inferred inheritance pattern, while 78 were no longer predicted to be at risk. *A priori* carrier risks were redefined with near- certainty (approaching 100%) in four fathers and five mothers, ∼50% in 59 second-degree relatives, and ∼25% in 51 third-degree relatives (Figure 1).

**Figure 1 A and B.**
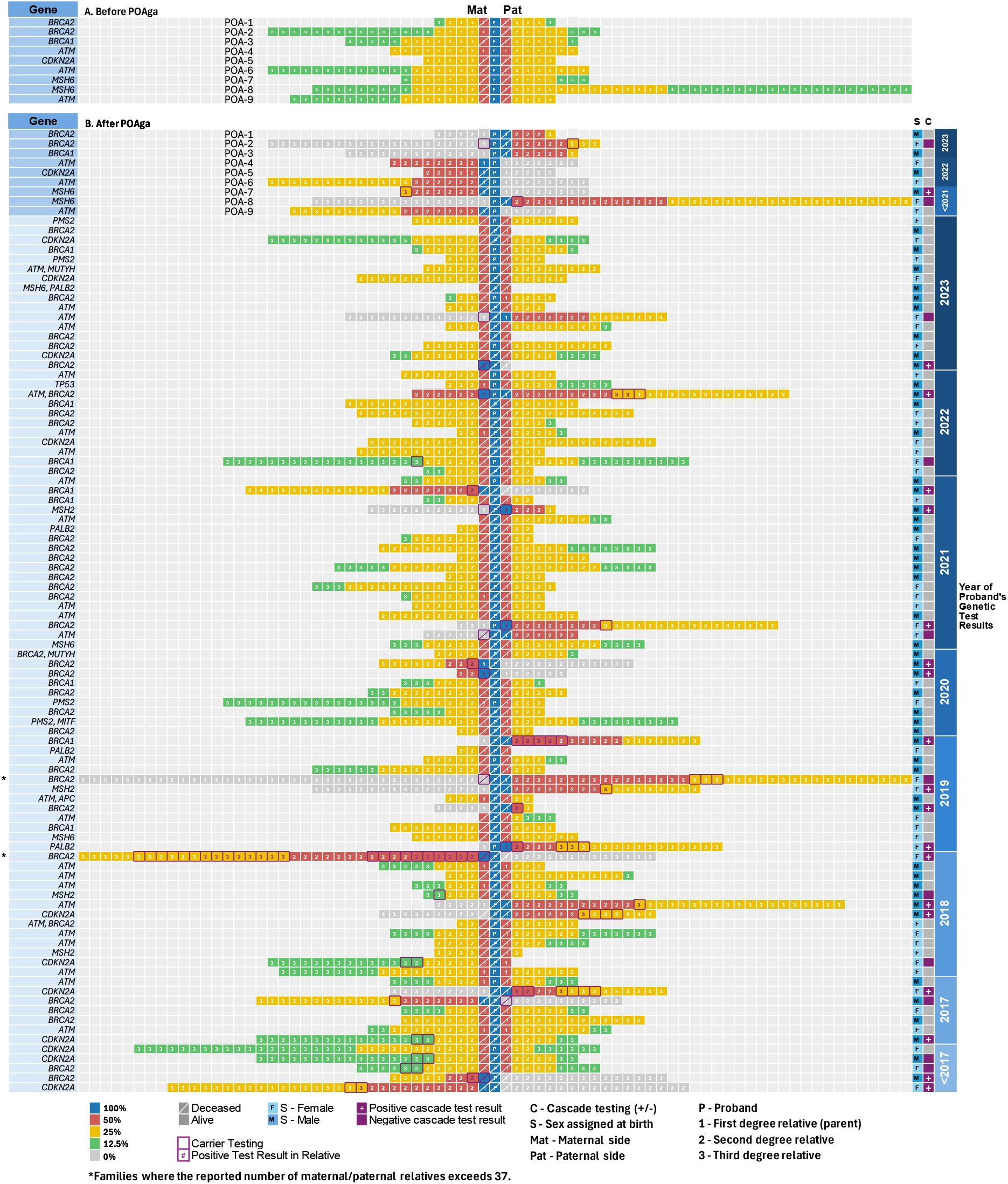
. Parent-of-Origin-Aware genomic analysis (POAga) enables refinement of risk estimates by inferring the parental origin of the variant, allowing for prioritization of cascade genetic testing towards the side of the family at highest risk. Predicted carrier risk estimates across 9 families with a known pathogenic variant where samples were available for POAga are shown before POAga (A) and after POAga (B), where the 9 probands who underwent POAga are listed at the top with the POAga informed risk assessment for the predicted side of the family, followed by 91 probands who did not undergo POAga. Numbered boxes denote individual family members. Numbers within denote the degree of relationship to the proband (P = proband, 1 = first-degree, 2 = second-degree, 3 = third-degree). Note, children and siblings are not included but proband’s half-siblings are included. Maternal relatives are to the left of the proband (P) and paternal relatives are to the right of the proband. Colored boxes indicate predicted carrier risk for the familial variant using: 100% (dark blue), 50% (red), 25% (yellow), 12.5% (green), and 0% (dark gray). Vital status for proband and parents is indicated (diagonal white line = deceased). Boxes with diagonal lines represent deceased probands or parents. Only vital status for proband and parents is indicated (diagonal white line = deceased). Cascade testing is indicated by a purple border; positive status for familial variant is indicated by purple font.

## Discussion

Parental segregation achieved without the need for parental samples could immediately impact genetic counselling practice and risk assessment of extended relatives. By determining the side of the family at-risk, POAga can increase the effectiveness and efficiency of risk communication in families while also providing clarity, and reducing uncertainty, anxiety and burden on patients and extended family members. While traditional approaches to cascade testing over the 10-year period were able to segregate the PV to the maternal or paternal lineage in one-fifth of families, POAga stands to improve this nearly 5-fold and provides the risk information immediately.

The fundamental alteration of familial risk assessment for these nine probands potentially relieved 78 individuals of the burden of genetic assessment and reclassified 59 second- degree relatives as having ∼50% risks, comparable to those traditionally ascribed to first- degree relatives. Considering the genes involved, their associated cancer risks, and existing screening guidelines, a 50% chance of carrying a PV/LPV in a known breast cancer susceptibility gene like *BRCA1* is considered medically actionable even before genetic status is confirmed, warranting eligibility for high-resolution breast imaging.(10) Hence, the application of germline testing and POAga in a PDAC population has precision oncology management implications for not only the patient but extending to their second-degree relatives.

Considering the 1192 extended relatives across the entire cohort who may have benefited from POAga, ensuring samples are collected from patients in a manner that may allow for secondary POAga will be critical. As such, we are exploring collection and storage of viable blood samples at the time of mainstreamed testing so that patients and families may access POAga if needed. Future prospective analysis will give insights into the impact of POAga on cascade testing rates.

As germline testing criteria expand and more older patients with cancer become eligible for testing, we anticipate that unresolved parental origin will become an increasingly important limitation to realizing the full preventive value of hereditary cancer identification. We envision this application of POAga as part of a broader public health approach to genomics that optimizes resource allocation and enables timely delivery of genetic counselling and hereditary cancer care to individuals most likely to benefit. Furthermore, as POAga already transforms the genetic risk assessment for patients and families by determining the parent of origin of a single autosomal variant, its broader ability to segregate all autosomal genomic variation is expected to further advance our understanding of hereditary cancer syndromes.

## Data Availability

Aggregate, de-identified data supporting the findings are included. Additional de-identified data may be available upon reasonable request, subject to institutional review and a data use agreement. No public secondary datasets were used.

## Acknowledgements and funding support

This work was supported by The Bob and Judy Hager Family Fund, Diamond Foundation, Genome Canada, Genome British Columbia, Canadian Institutes of Health Research, BC Cancer Foundation, and Vancouver General Hospital Foundation. Carol Cremin acknowledges support from Michael Smith Health Research BC through a Health Professional-Investigator Award.

## References

1. Wang Y, Golesworthy B, Cuggia A, Domecq C, Chaudhury P, Barkun J, et al. Oncology clinic-based germline genetic testing for exocrine pancreatic cancer enables timely return of results and unveils low uptake of cascade testing. J Med Genet. 2022 Aug;59(8):793–800.

2. Blandy C, Chabal F, Stoppa-Lyonnet D, Julian-Reynier C. Testing participation in BRCA1/2-positive families: initiator role of index cases. Genet Test. 2003 Sep;7(3):225–33.

3. Fehniger J, Lin F, Beattie MS, Joseph G, Kaplan C. Family communication of BRCA1/2 results and family uptake of BRCA1/2 testing in a diverse population of BRCA1/2 carriers. J Genet Couns. 2013 Oct;22(5):603–12.

4. Frey MK, Ahsan MD, Bergeron H, Lin J, Li X, Fowlkes RK, et al. Cascade testing for hereditary cancer syndromes: should we move toward direct relative contact? A systematic review and meta-analysis. J Clin Oncol. 2022 Dec 10;40(35):4129–43.

5. Tiller J, Otlowski M. Maximizing cascade genetic testing for disease prevention through direct notification of at-risk relatives. Nat Med. 2025 Oct;31(10):3225–6.

6. Chiang J, Chua Z, Chan JY, Sule AA, Loke WH, Lum E, et al. Strategies to improve implementation of cascade testing in hereditary cancer syndromes: a systematic review. NPJ Genom Med. 2024 Apr 3;9(1):26.

7. Uson PLS Jr, Samadder NJ, Riegert-Johnson D, Boardman L, Borad MJ, Ahn D, et al. Clinical impact of pathogenic germline variants in pancreatic cancer: results from a multicenter, prospective, universal genetic testing study. Clin Transl Gastroenterol. 2021 Oct 8;12(10):e00414.

8. Jacobs MF, Stoffel EM. Genetic and other risk factors for pancreatic ductal adenocarcinoma (PDAC). Fam Cancer. 2024 Aug;23(3):221–32.

9. Akbari V, Hanlon VCT, O’Neill K, Lefebvre L, Schrader KA, Lansdorp PM, et al. Parent-of-origin detection and chromosome-scale haplotyping using long-read DNA methylation sequencing and Strand-seq. Cell Genom. 2022 Dec 21;3(1):100233.

10. Lubinski J, Kotsopoulos J, Moller P, Pal T, Eisen A, Peck L, et al. MRI surveillance and breast cancer mortality in women with BRCA1 and BRCA2 sequence variations. JAMA Oncol. 2024 Apr;10(4):493–9.

